# Cognitive Costs and Gait Parameters During Single- and Dual-Task Conditions: A Comparative Study in Individuals With and Without Non-Specific Neck Pain

**DOI:** 10.1101/2025.01.28.25321245

**Authors:** Ibrahim M. Moustafa, Shorouk Abu-Ghosh, Amal Ahbouch, Shima Abdollah Mohammad Zadeh, Meeyoung Kim, Iman Khowailed

## Abstract

**Background:** The high incidence of non-specific neck pain (NSNP) among students, commonly associated with prolonged sedentary behavior and extensive use of electronic devices, highlights the need to assess its impact on cognitive and motor functions. This study aims to evaluate the effects of NSNP on dual-task performance by analyzing gait parameters and cognitive performance during both single-task and dual-task conditions.

**Methods:** Forty-five participants with NSNP and forty-five age-matched controls were assessed using an optical motion-capture system. Participants underwent gait assessments during both single-task (without cognitive load) and dual-task conditions, in which the cognitive tasks involved simple mathematical computations.

**Results:** Results revealed that, under single-task conditions, differences in gait parameters between groups were not statistically significant. However, under dual-task conditions, participants with NSNP exhibited significant impairments in gait parameters and higher cognitive costs (p < 0.05). Correlation analysis indicated that pain intensity was significantly associated with cognitive cost and gait parameter alterations during dual-task conditions (p < 0.05).

**Conclusions:** These findings suggest that NSNP significantly elevates the cognitive effort required during dual tasking. These finding emphasizes on the need for interventions to alleviate neck pain and improve both physical and cognitive health.

## 1. Introduction

Neck pain has become increasingly prevalent among students, mainly due to prolonged periods of studying, sedentary behavior, and extensive use of electronic devices [1]. As students spend more time hunched over books and screens, their necks bear the burden of the strain [2]. This non-specific neck pain (NSNP), which is not linked to any specific pathology or anatomical defect, affects a significant number of students and can lead to considerable discomfort, impacting not only their academic performance but also their overall quality of life [3].

The prevalence of neck pain among students highlights the importance of understanding its causes and effects. Factors such as poor posture, lack of ergonomic workstations, and inadequate physical activity contribute to the development of this condition [4]. Evidence has shown that improper use of pillows, prolonged use of electronic devices, and poor sitting posture are significant risk factors for neck pain in this population [5]. Moreover, neck pain is influenced by various modifiable and non-modifiable risk factors, such as advanced age, female gender, low social support, and a history of neck or lower back pain, emphasizing the need for prevention and early diagnosis [6].

It is crucial for students to take proactive steps to alleviate neck pain, such as practicing proper posture, taking regular breaks, incorporating neck stretches into their daily routines, and seeking medical attention if necessary [7]. By addressing neck pain and prioritizing their well-being, students can enhance their focus, productivity, and overall health[8,9]. Educational institutions can also play a role by promoting ergonomic practices and providing resources for physical well-being [10].

While the physical symptoms of NSNP are well-documented, there is a significant and noteworthy surge in interest surrounding the comprehensive understanding of its cognitive implications. It is undeniably established that chronic pain, including neck pain, can profoundly and unequivocally influence cognitive function through an intricate network of mechanisms[11]. These mechanisms encompass pain-induced distraction, extensively altered sensory input, and the overwhelming psychological stress that is inherently intertwined with the chronically discomforting nature of this condition [12]. These multifaceted and interrelated factors collectively and synergistically impose a substantial cognitive burden upon individuals, rendering the performance of tasks that necessitate concurrent physical and cognitive effort all the more arduous and formidable [13].

Literature has shown that chronic pain conditions can impair cognitive performance, particularly in tasks requiring attention, memory, and executive function [14]. However, it remains to be seen how mild to moderate neck pain among students affects their gait parameters, especially under dual-task conditions. The dual-task paradigm, which involves performing a cognitive task while simultaneously engaging in a physical activity, is particularly useful for studying the influence of cognitive load on motor performance [15].

Understanding dual-task performance can provide essentials insights into the impact of neck pain on individuals’ ability to effectively manage multiple concurrent tasks [16]. This is particularly crucial for various daily activities where cognitive-motor integration is essential [17]. For instance, students frequently find themselves needing to engage in conversations while walking or engaging in mental calculations while in motion. These situations necessitate seamless coordination between cognitive and motor functions [18]. However, for individuals experiencing neck pain, the additional burden of discomfort and pain may intensify the challenges associated with maintaining equilibrium and coordination, potentially resulting in compromised performance for both tasks at hand [19].

Therefore, it is crucial to understand how neck pain affects dual-task performance to fully grasp the overall impact of this condition. Additionally, by exploring the interplay between physical discomfort and cognitive load, we can better appreciate the multifaceted nature of neck pain and its broader implications. The purpose of this study is to investigate the effects of NSNP on dual-task performance by examining gait parameters and cognitive performance during single-task and dual-task conditions among students. By comparing individuals with and without neck pain, we seek to identify the cognitive costs associated with managing dual tasks in the presence of neck pain. Ultimately, our goal is to promote a holistic approach to student well-being, recognizing the interconnectedness of physical and mental health in achieving optimal academic and personal outcomes.

## 2. Materials and Methods

### 2.1. Study Design, Participants and Setting

This comparative study was designed to assess the impact of cognitive dual-tasking on gait parameters in students experiencing from NSNP, in comparison to a well-matched control group without neck pain. The research was conducted in the laboratories of the University of Sharjah, with subjects recruited from among the university’s students and staff through social media outreach. Recruitment took place from January to May 2024.

### 2.2. Ethical Considerations

The study received ethical approval from the University of Sharjah’s College of Health Sciences (Ethical approval number: REC-23-05-11-03-S). All participants provided their informed written consent prior to the start of data collection. The study adhered to established ethical guidelines and regulations

### 2.3. Inclusion and exclusion criteria

- Participants were eligible for the NSNP group if they met the following criteria:
  1. Age between 18 and 24 years;
  2. Experience of NSNP for at least three months, with a pain intensity rating between 30 and 70 on the Visual Analog Scale (VAS);
  3. Affiliation as a student at the University of Sharjah;
  4. Willingness to participate in the study and provide informed consent. The control group consisted of individuals within the same age range and affiliation who reported no neck pain.
- Participants were excluded from the study if they had:
  1. Previous fractures or operations on the neck or shoulders;
  2. Previous major injuries or surgical procedures involving the musculoskeletal system;
  3. Diagnosis of fibromyalgia, cervical radiculopathy/myelopathy, or cognitive impairments;
  4. Deformities in the spine or limbs. These criteria ensured a homogeneous sample, reducing potential confounding factors that could influence gait or cognitive performance outcomes.

### 2.3. Study Tools and Outcome Measures

#### 2.3.1. Numerical Pain Rating Scale

Participants quantified their neck pain intensity using a numerical pain rating scale, a validated and reliable instrument for measuring pain intensity.

#### 2.3.2. BTS GAITLAB System

The spatiotemporal and kinematic parameters of gait were assessed using an optical motion-capture system consisting of 8 infrared cameras (Smart-D, BTS Bioengineering, Milan, Italy) running at a frequency of 120 Hz. Anthropometric measurements, such as height, weight, and various body segment widths, were obtained before the experimental tests. Spherical reflective passive markers were placed on the subjects’ skin following the protocol outlined by Davis et al.

#### 2.3.3. Dual Task Cost (DTC) Percentage

The analysis was based on average performance from three dual-task trials to establish a comprehensive measure of the impact of cognitive load on gait. To calculate the Dual Task Cost (DTC) percentage, the disparity between single and dual-task performance scores was determined, and then divided by the single-task performance score before being multiplied by 100. For instance, if the average speed during single-task walking was 1.2 m/s and during dual-task walking was 1.0 m/s, the DTC percentage would be calculated as ((1.2 -1.0) / 1.2) × 100 = 16.67%.

#### 2.3.4. Spatiotemporal Parameters

The spatiotemporal parameters of the participants, including step length, speed, and cadence, were measured under both single-task and dual-task conditions using the BTS gait lab system

### 2.4. Study Procedure

Participants were asked to walk at their chosen pace along a 10-meter walkway, with their 3D marker trajectories recorded by cameras under two different scenarios: single-task and dual-task. In the single-task scenario, participants walked at their chosen speed without performing any additional tasks, completing three trials to establish their baseline gait characteristics. In the dual-task scenario, participants walked while engaging in various cognitive tasks, such as answering yes or no questions, counting, or performing simple math calculations. Each participant completed a minimum of three trials for each scenario. Upon completion of the tests, the raw data were analyzed using specialized software (Smart Analyzer, BTS Bioengineering, Milan, Italy) to derive the necessary parameters.

### 2.5. Sample Size Determination

The sample size was determined based on an effect size of 0.5, a two-tailed significance level (α) of 0.05, and a statistical power (1−β) of 0.80. Using G*Power 3.1 software, the calculation indicated that a minimum of 40 participants per group (neck pain and no neck pain) would be required to detect a significant difference with 80% power at a 5% significance level. To accommodate potential dropouts, 5 additional participants were included per group, bringing the total to 45 participants per group.

### 2.6. Statistical Analysis

To assess the normal distribution of all baseline descriptive variables, the Kolmogorov-Smirnov test was utilized. Continuous variables are presented as mean values alongside their standard deviations (SD). Levene’s test was applied at a 95% confidence interval to examine the homogeneity of variances, with significance defined as p-values less than 0.05. Descriptive statistics (mean ± SD) were reported for each respective time point. To confirm group equivalence, chi-squared tests were applied to categorical variables, while Student’s t-tests were used for continuous variables. The comparison of mean values between groups was conducted using the Student’s t-test, with statistical significance established at a p-value of less than 0.05. Effect size was assessed using Cohen’s d, where d values around 0.2, 0.5, and 0.8 corresponded to negligible, moderate, and substantial clinical significance, respectively. Pearson’s correlation coefficients (r) were utilized to investigate the associations between neck pain severity and spatiotemporal gait parameters in both single and dual-task conditions, as well as the relationship between neck pain severity and cognitive cost. All analyses were performed using SPSS version 20.0 (SPSS Inc., Chicago, IL, USA), ensuring the assumptions of normality and homogeneity of variance were met prior to analysis.

## 3. Results

### 3.1. Demographics

Over 200 potential participants were initially screened, with the most common exclusion criterion being the pain levels outside 30-70 on the VAS or having neck pain for less than three months. Ultimately, 45 participants with NSNP (25 males, 20 females) and 45 matched controls (based on age, BMI, and sex) without neck pain were recruited for the study (p>0.05). The significant difference in VAS validates the classification of the groups based on neck pain status. The demographic characteristics and clinical variables of these participants are detailed in Table 1.

**Table 1.**
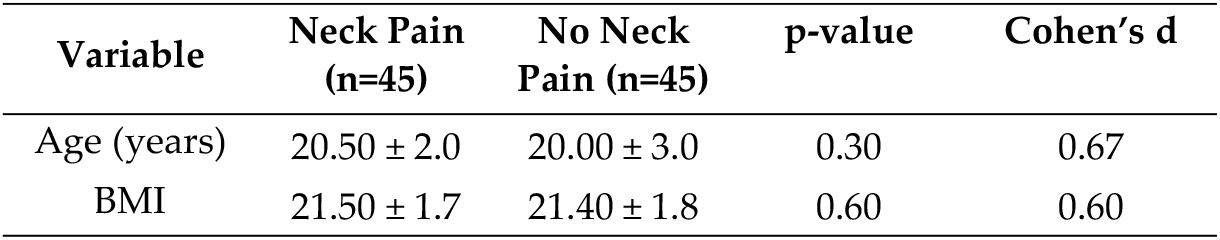
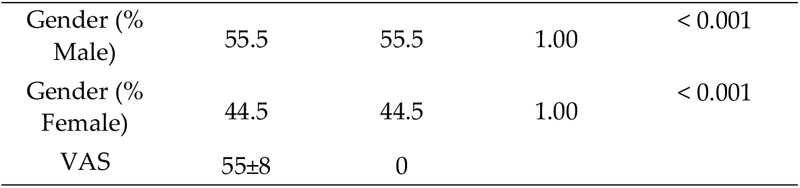
Baseline demographic characteristics and clinical variables.

### 3.2. Single Task Detailed Results

Table 2 presents the mean differences in spatiotemporal parameters between the neck pain and no neck pain groups during the single-task condition. Most variables exhibit non-significant differences. Most spatiotemporal parameters during the single-task condition do not show significant differences between the neck pain and no neck pain groups. However, significant differences were observed in swing time, mean velocity and step length, indicating an impact of neck pain on these specific parameters.

**Table 2.** Mean Differences between Spatiotemporal Parameters in Neck Pain and No Neck Pain Groups during Single Task.

| Variable | Group | Mean ± SD | 95% CI | Cohen's d | p-value |
| --- | --- | --- | --- | --- | --- |
| Stride time (s) | Neck Pain | 1.25 ± 0.10 | [1.22, 1.28] | 0.20 | 0.15 |
|  | No Neck Pain | 1.23 ± 0.10 | [1.20, 1.26] |  |  |
| Stance time (s) | Neck Pain | 0.75 ± 0.05 | [0.73, 0.77] | 0.25 | 0.10 |
|  | No Neck Pain | 0.74 ± 0.05 | [0.72, 0.76] |  |  |
| Swing time (s) | Neck Pain | 0.45 ± 0.05 | [0.43, 0.47] | 0.60 | < 0.001 |
|  | No Neck Pain | 0.42 ± 0.05 | [0.40, 0.44] |  |  |
| Stance phase (%) | Neck Pain | 62.0 ± 2.0 | [61.3, 62.7] | 0.30 | 0.05 |
|  | No Neck Pain | 61.5 ± 2.0 | [60.8, 62.2] |  |  |
| Double support phase (%) | Neck Pain | 12.0 ± 1.5 | [11.5, 12.5] | 0.15 | 0.20 |
|  | No Neck Pain | 11.8 ± 1.5 | [11.3, 12.3] |  |  |
| Mean velocity m/s | Neck Pain | 1.10 ± 0.10 | [1.07, 1.13] | 0.40 | 0.02 |
|  | No Neck Pain | 1.15 ± 0.10 | [1.12, 1.18] |  |  |
| Mean velocity height (%height/s) | Neck Pain | 70.0 ± 5.0 | [68.5, 71.5] | 0.10 | 0.50 |
|  | No Neck Pain | 69.8 ± 5.0 | [68.3, 71.3] |  |  |
| Cadence (steps/min) | Neck Pain | 110.0 ± 5.0 | [108.5, 111.5] | 0.20 | 0.15 |
|  | No Neck Pain | 109.0 ± 5.0 | [107.5, 110.5] |  |  |
| Stride length (%height) | Neck Pain | 75.0 ± 5.0 | [73.5, 76.5] | 0.15 | 0.20 |
|  | No Neck Pain | 74.5 ± 5.0 | [73.0, 76.0] |  |  |
| Step length (m) | Neck Pain | 0.60 ± 0.05 | [0.58, 0.62] | 0.60 | < 0.001 |
|  | No Neck Pain | 0.57 ± 0.05 | [0.55, 0.59] |  |  |
| Step width avg (m) | Neck Pain | 0.09 ± 0.03 | [0.08, 0.10] | 0.10 | 0.50 |
|  | No Neck Pain | 0.09 ± 0.03 | [0.08, 0.10] |  |  |

### 3.3. Dual Task Detailed Results

Table 3 illustrates the mean differences in spatiotemporal parameters between the neck pain and no neck pain groups during the dual-task condition. Significant differences were observed in all spatiotemporal parameters, except for the cadence, between the neck pain and no neck pain groups during the dual-task condition. This indicates that neck pain significantly impacts gait parameters under increased cognitive load.

**Table 3.** Mean Differences in Spatiotemporal Parameters Between Neck Pain and No Neck Pain Groups During Dual-Task Conditions.

| Variable | Group | Mean ± SD | 95% CI | Cohen's d | p-value |
| --- | --- | --- | --- | --- | --- |
| Stride time (s) | Neck Pain | 1.50 ± 0.15 | [1.45, 1.55] | 0.80 | < 0.001 |
| | No Neck Pain | 1.20 ± 0.10 | [1.17, 1.23] | | |
| Stance time (s) | Neck Pain | 0.80 ± 0.05 | [0.78, 0.82] | 1.14 | < 0.001 |
| | No Neck Pain | 0.70 ± 0.05 | [0.68, 0.72] | | |
| Swing time (s) | Neck Pain | 0.50 ± 0.05 | [0.48, 0.52] | 0.80 | < 0.001 |
| | No Neck Pain | 0.45 ± 0.05 | [0.43, 0.47] | | |
| Stance phase (%) | Neck Pain | 65.0 ± 3.0 | [64.1, 65.9] | 0.80 | 0.01 |
| | No Neck Pain | 61.0 ± 2.5 | [60.3, 61.7] | | |
| Double support phase (%) | Neck Pain | 13.0 ± 2.0 | [12.4, 13.6] | 0.80 | < 0.001 |
| | No Neck Pain | 11.0 ± 1.5 | [10.5, 11.5] | | |
| Mean velocity m/s | Neck Pain | 1.10 ± 0.10 | [1.07, 1.13] | 0.80 | < 0.001 |
| | No Neck Pain | 1.20 ± 0.10 | [1.17, 1.23] | | |
| Mean velocity height (%height/s) | Neck Pain | 65.0 ± 5.0 | [63.5, 66.5] | 0.80 | < 0.001 |
| | No Neck Pain | 72.0 ± 5.0 | [70.5, 73.5] | | |
| Cadence (steps/min) | Neck Pain | 110.0 ± 5.0 | [108.5, 111.5] | 0.60 | 0.30 |
| | No Neck Pain | 105.0 ± 5.0 | [103.5, 106.5] | | |
| Stride length (%height) | Neck Pain | 75.0 ± 5.0 | [73.5, 76.5] | 1.00 | < 0.001 |
| | No Neck Pain | 80.0 ± 5.0 | [78.5, 81.5] | | |
| Step length (m) | Neck Pain | 0.60 ± 0.05 | [0.58, 0.62] | 1.10 | < 0.001 |
| | No Neck Pain | 0.65 ± 0.05 | [0.63, 0.67] | | |
| Step width avg (m) | Neck Pain | 0.09 ± 0.03 | [0.08, 0.10] | 0.60 | < 0.001 |
| | No Neck Pain | 0.07 ± 0.02 | [0.06, 0.08] | | |

### 3.4. Cognitive Cost Detailed Results

The cognitive cost comparison results, presented in Table 4, show significant differences across multiple gait parameters in the neck pain group. The Mean Difference in Cognitive Cost (CC) between Neck Pain and No Neck Pain. The cognitive cost values indicate that individuals with neck pain incur significantly higher cognitive costs across various gait parameters during dual-task conditions compared to those without neck pain. This finding suggests that neck pain exacerbates the cognitive effort required to maintain gait.

**Table 4.** Mean Differences in cognitive cost Between Neck Pain and No Neck Pain groups.

| Variable | Group | Mean ± SD | 95% CI | Cohen's d | p-value |
| --- | --- | --- | --- | --- | --- |
| CC Stride time | Neck Pain | 25.79 ± 5.95 | [23.99, 27.59] | 0.87 | < 0.001 |
|  | No Neck Pain | 5.19 ± 3.50 | [4.19, 6.19] |  |  |
| CC Stance time | Neck Pain | 8.43 ± 4.00 | [7.43, 9.43] | 0.89 | < 0.001 |
|  | No Neck Pain | -6.75 ± 3.75 | [-7.75, -5.75] |  |  |
| CC Swing time | Neck Pain | 2.78 ± 3.00 | [2.08, 3.48] | -0.72 | < 0.001 |
|  | No Neck Pain | 15.03 ± 4.68 | [13.53, 16.53] |  |  |
| CC Stance phase | Neck Pain | 8.49 ± 3.19 | [7.49, 9.49] | 0.64 | 0.01 |
|  | No Neck Pain | 1.40 ± 3.00 | [0.70, 2.10] |  |  |
| CC Double support phase | Neck Pain | 19.87 ± 5.69 | [18.07, 21.67] | 0.66 | 0.01 |
|  | No Neck Pain | 1.11 ± 5.00 | [-0.39, 2.61] |  |  |
| CC Mean velocity | Neck Pain | 12.36 ± 6.45 | [10.36, 14.36] | -0.75 | 0.02 |
|  | No Neck Pain | 6.85 ± 4.00 | [5.85, 7.85] |  |  |
| CC Mean velocity height | Neck Pain | 13.34 ± 4.90 | [11.84, 14.84] | -0.80 | < 0.001 |
|  | No Neck Pain | 3.18 ± 4.00 | [2.18, 4.18] |  |  |
| CC Cadence | Neck Pain | 4.49 ± 4.00 | [3.49, 5.49] | -0.30 | < 0.001 |
|  | No Neck Pain | 1.46 ± 4.00 | [0.46, 2.46] |  |  |
| CC Stride length | Neck Pain | 6.84 ± 4.00 | [5.84, 7.84] | -0.81 | 0.01 |
|  | No Neck Pain | 2.41 ± 4.00 | [1.41, 3.41] |  |  |
| CC Step length | Neck Pain | 9.98 ± 5.00 | [8.48, 11.48] | -1.13 | 0.01 |
|  | No Neck Pain | 4.39 ± 4.00 | [3.39, 5.39] |  |  |
| CC Step width avg | Neck Pain | 12.43 ± 4.90 | [10.93, 13.93] | 0.49 | 0.01 |
|  | No Neck Pain | 9.91 ± 5.00 | [8.41, 11.41] |  |  |

### 3.5. Correlational Results

The single-task condition reveals no significant correlations between pain intensity and gait parameters (p > 0.05). This indicates that pain intensity does not significantly influence performance parameters during single tasks in either group.

The correlation table for the dual-task condition illustrates significant correlations between pain intensity and various performance parameters, suggesting that pain intensity substantially influences gait parameters during dual-task activities (Table 5).

**Table 5.**
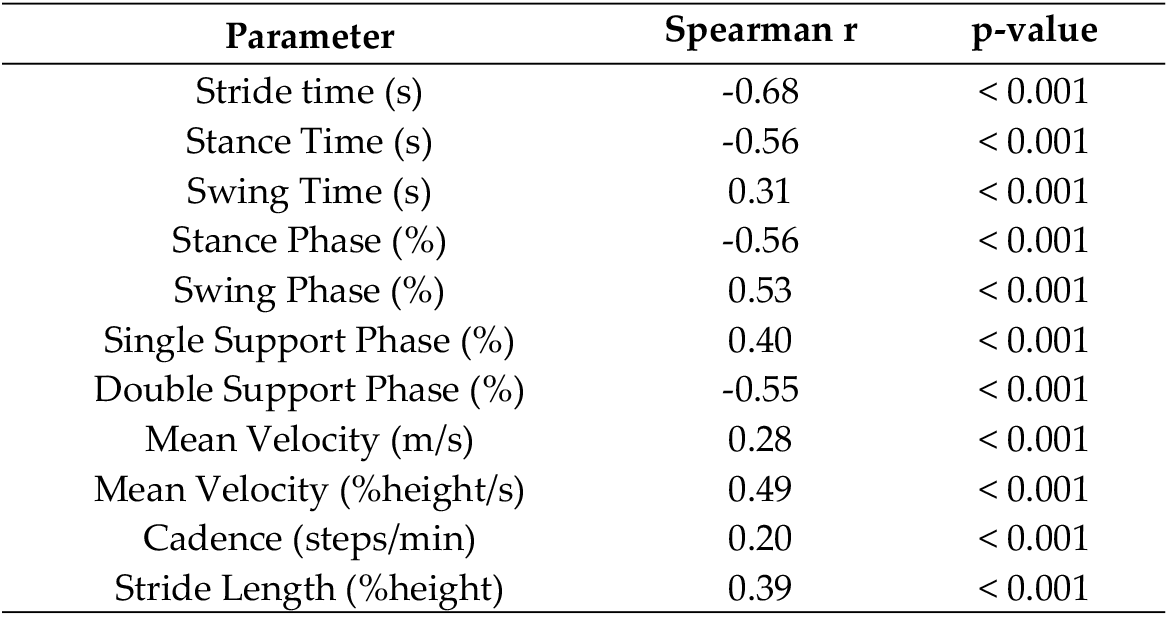
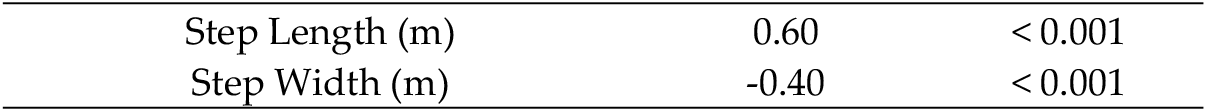
Correlation Between Pain Intensity and Gait Parameters During Dual-Task Conditions.

The correlation table for cognitive cost values illustrates a connection between pain intensity and cognitive cost measures (Table 6). The results suggest that pain intensity significantly influences the cognitive cost related to various gait parameters, indicating that as pain intensity rises, the cognitive effort needed to sustain gait during dual-task scenarios also increases.

**Table 6.** Correlation Between Pain Intensity and Cognitive Cost.

| Parameter | Spearman r | p-value |
| --- | --- | --- |
| CC Stride time (s) | -0.61 | < 0.001 |
| CC Stance Time (s) | -0.41 | < 0.001 |
| CC Swing Time (s) | -0.35 | < 0.001 |
| CC Stance Phase (%) | -0.35 | < 0.001 |
| CC Swing Phase (%) | -0.45 | < 0.001 |
| CC Single Support Phase (%) | -0.23 | 0.04 |
| CC Double Support Phase (%) | -0.21 | 0.04 |
| CC Mean Velocity (m/s) | -0.37 | 0.01 |
| CC Mean Velocity (%height/s) | -0.30 | 0.01 |
| CC Cadence (steps/min) | -0.22 | 0.01 |
| CC Stride Length (%height) | -0.32 | 0.01 |
| CC Step Length (m) | -0.55 | < 0.001 |
| CC Step Width (m) | -0.50 | < 0.001 |

CC: Cognitive cost

## 4. Discussion

Our study showed that how NSNP influences cognitive performance during dual-task activities, bringing forth crucial findings that deepen our understanding of the connection between musculoskeletal discomfort and cognitive function. The data demonstrate that individuals with NSNP incur higher cognitive costs when performing dual tasks compared to those without neck pain, indicating that NSNP might elevate cognitive demands during gait-related activities. These outcomes are in line with prior research, which has shown that neck pain can lead to both physiological and neurological disruptions, thereby affecting physical and cognitive functions. [20].

These findings enhance our grasping the intricate nature of neck pain and its wider implications. This observation aligns with cognitive load theory, which argues that physical discomfort and proprioceptive errors linked to neck pain can raise cognitive load, thereby negatively affecting cognitive performance. Recent studies reinforce this idea by showing that neck pain disrupts sensorimotor control and neural processing, leading to an increased cognitive burden and reduced efficiency in cognitive tasks.[21]

### 4.1. Interpretation of Findings

The discrepancies in dual-task performance between the neck pain group and the control group are in line with prior studies that indicate musculoskeletal discomfort can influence cognitive processes [22–24]. The increased cognitive load observed in the neck pain group may result from disrupted sensory input and proprioception caused by pain and discomfort [25–27]. This phenomenon is like phantom limb pain, which emerges from maladaptive cortical reorganization after sensory input loss, suggesting that NSNP might also trigger maladaptive changes in the sensory and motor cortices [28,29]. This is further supported by research showing that both conditions involve reorganization within the primary somatosensory and motor cortices, which contributes to chronic pain [30,31]. These disturbances may impose an increased cognitive load as the brain compensates for compromised postural stability. It is a phenomenon elucidated by prior research which underscores deficits in attention, processing speed, and memory among individuals experiencing musculoskeletal pain. [32].

Several mechanisms may account for the increased cognitive burden observed in the NSNP group. First, neck pain creates biomechanical stress on the neck and shoulders, leading to muscle fatigue and discomfort, which can reduce the cognitive resources available for task execution [6]. Additionally, neck pain is linked to altered proprioceptive function, which can disrupt sensory integration and further increase the cognitive load needed to maintain balance and perform tasks [33]. Structural changes in the cervical spine associated with neck pain may also impair cerebral blood flow, affecting cognitive functions such as attention and memory [34,35]. Deafferentation due to poor posture can intensify these cognitive deficits, as research shows that the loss of sensory nerve signals from poor posture increases cognitive load by disrupting proprioceptive feedback and amplifying the brain’s compensatory efforts [36,37].

Additionally, neuroplasticity plays a pivotal role in the interplay between cognitive processes and musculoskeletal discomfort [22]. Neuroplastic changes induced by altered sensory input from neck pain can result in both beneficial and adverse cognitive outcomes [38]. Studies indicate that neuroplastic mechanisms, such as synaptogenesis and cortical reorganization, are engaged to accommodate the elevated cognitive demands imposed by neck pain. These processes highlight the brain’s profound capacity to reorganize and recalibrate its neural networks in response to altered sensory inputs, thereby preserving cognitive function despite the presence of physical impairments [38,39].

### 4.2. Implications for Clinical Practice

The robust correlations between pain intensity and cognitive dual-task performance emphasize the necessity for comprehensive assessment protocols that address both musculoskeletal and cognitive dimensions. Clinicians should recognize the potential cognitive effects of neck pain and incorporate strategies to manage musculoskeletal discomfort within their rehabilitation approaches. [40]. This is particularly crucial for populations at higher risk of cognitive decline, including older adults and individuals with neurological disorders [41,42]. Our research also highlights the importance of dual-task gait assessments as a crucial tool for identifying cognitive impairments. By integrating cognitive and motor tasks, these assessments provide a more realistic measure of everyday functioning, making them essential for detecting subtle cognitive deficits that may not be evident in single-task evaluations. [43]. These findings have significant implications for early intervention and continuous monitoring of cognitive health in clinical practice. Clinicians should incorporate neck pain assessments into routine evaluations, especially for individuals exhibiting cognitive concerns. [44]. Embracing interdisciplinary approaches involving physical therapists, occupational therapists, and cognitive specialists can provide comprehensive care that addresses both the physical and cognitive challenges associated with neck pain [45]. Early intervention and patient education emphasizing the importance of proper posture and effective management of musculoskeletal discomfort are essential for mitigating the risk of developing neck pain and its consequent cognitive impairments [46].

### 4.3. Practical Applications

People suffering from neck pain are more likely to experience increased cognitive fatigue and a decline in efficiency when performing tasks that require both mental and physical effort simultaneously [47]. This can greatly impact everyday activities, work productivity, and overall quality of life [48]. Additionally, chronic neck pain can lead to enduring health problems, such as persistent pain, headaches, and even potential cognitive decline over time [49–51].

To counteract the negative effects of neck pain, it is vital to implement strategies that alleviate discomfort. Such strategies might include physical therapy, workplace ergonomic modifications, and exercises designed to strengthen the neck and upper back muscles [52]. Furthermore, taking regular breaks and performing posture checks during extended computer use can help maintain proper alignment and reduce the risk of developing neck pain [53].

## Limitations and Future Directions

Although our study is well-designed and yields significant findings, it is important to recognize certain limitations. The cross-sectional design restricts our ability to infer causality, emphasizing the need for longitudinal studies to better understand the temporal relationship between neck pain and cognitive decline. Additionally, our sample consisted primarily of university students and staff, which may not fully represent the broader population. Future studies should aim to include more diverse participant groups to improve the generalizability of the results.

Further research is necessary to explore the mechanisms connecting musculoskeletal discomfort with cognitive function. Specifically, examining the roles and connectivity of different brain regions in individuals experiencing musculoskeletal pain could shed light on the neurophysiological underpinnings of our findings. Moreover, intervention studies that investigate the impact of pain relief on cognitive outcomes will be critical for evaluating the effectiveness of targeted treatments.

## 5. Conclusions

Our research indicates that NSNP markedly raises the cognitive demands of walking, emphasizing the urgent need to manage musculoskeletal discomfort to safeguard cognitive function. Implementing targeted strategies to alleviate neck pain could lead to improvements in both physical and mental health, thereby enhancing overall quality of life and daily activities. Additional studies are necessary to investigate the long-term consequences of neck pain and to determine the effectiveness of different interventions in minimizing its cognitive effects.

## Data Availability Statement

Data is provided as supplementary information file.

